# Describing the COVID-19 Outbreak Fitting Modified SIR models to Data

**DOI:** 10.1101/2020.04.29.20084285

**Authors:** Aldo Ianni, Nicola Rossi

## Abstract

In this paper we fit simple modifications of the SIR compartmental model to the COVID-19 outbreak data, available from official sources for Italy and other countries. Even if the complexity of the pandemic can not be easily modelled, we show that our model, at present, describes the time evolution of the data in spite of the application of the social distancing and lock-down procedure. Finally, we discuss the reliability of the model predictions, under certain conditions, for estimating the near and far future evolution of the COVID-19 outbreak. The conditions for the applicability of the proposed models are discussed.

## 1 Introduction

The Coronavirus Disease, named COVID-19 or SARS-Cov-2, is an infectious disease caused by a newly discovered coronavirus [1]. Most people infected with the COVID-19 virus experiences from mild to moderate respiratory illness and recover without requiring special treatment. Older people, and those with underlying preexisting medical problems like cardiovascular diseases, diabetes, chronic respiratory diseases, and cancer are more likely to develop serious illness. Yet, at present much needs to be understood about the effects of this new virus. COVID-19 spreads quickly, mainly through droplets of saliva or discharge from the nose when an infected person coughs or sneezes. The coronavirus in airborne droplets can survive for many tens of minutes and eventually sticks to surfaces and remains active up to many hours, depending on materials [2]. This fact could further increase the diffusion of the outbreak.

The velocity of the outbreak spread is described by the so-called basic reproduction number (or ratio), usually referred to as R_0_, defined as the expected number of new infections from a single new case in a population where all subjects are susceptible. The lock-down and/or the social distancing should help in reducing *R*_0_, making the outbreak duration shorter than the typical time needed by the free natural evolution until the so-call “herd immunity” is reached.

Another important aspect of COVID-19, and in general for outbreaks in progress, is the impossibility of quantifying the asymptomatic population. Many ongoing studies, as e.g. [3], are indeed showing that this component could be a substantial fraction of the visible cases, i.e. the ones detected by reliable medical tests. Even if not detected, the asymptomatic cases can play an important role in the diffusion of the coronavirus epidemic and can drastically change the outcome of the model predictions.

In this work we try to infer *R*_0_ and other parameters from data and to predict the evolution of the outbreak, analyzing simultaneously the cumulative behaviour of active and removed (namely “recovered+deceased”) cases.

In Sec. 2 we introduce a time dependence of the parameters of the standard SIR compartmental model. In Sec. 3 we introduce a change in the model to account for recovered and deceased compartments separately. In Sec. 4 we discuss results for Italian data. In Sec. 5 we discuss results for Spanish data as an example of application to other data-set. Finally, in Sec. 6 we present a model which tries to accounts for the asymptomatic hidden component, that is not yet detected for various practical reasons.

## 2 SIR-T: A Time Dependent SIR Model

### 2.1 From SIR to SIR-T

In order to study the spread of the COVID-19 we take into account the basic SIR model [4]. In the model we distinguish three categories of individuals, known as “compartments”: *S* for the number of susceptible, *I* for the number of infectious and *R* for the number of recovered. The last category might include recovered and deceased together or separately. Based on this assumption the mathematical model is written in terms of the following set of differential equations:

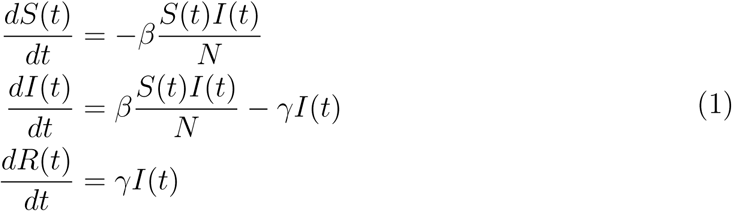

where *N* = *S*(*t*) + *I*(*t*) + *R*(*t*) is constant by definition being zero the sum of their derivatives, *β* determines the transmission rate and *γ* the recovery rate. If a susceptible population of (*S*(*t* = 0) = *N*) people is affected for the first time by the outbreak, the initial conditions of the system (1) can be easily cast as: *N*(0) = *N*_0_, *I*(0) = *I*_0_ and *R*(0) = 0.

In the SIR model the *R*_0_, determines the dynamic of the infection, and emerges as the ratio *β*/*γ*. With this definition we can write:

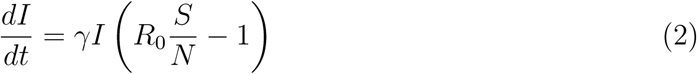

From eqs. (5) it yields that: 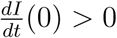 if *R*_0_ *> N/S*(0). Therefore, the epidemic decreases when *R*_0_ < *N*/*S*(0).

In our model we introduce a time dependence for *R*_0_ through the time dependence of the parameter *β*, to account for containment measures, such as population and commercial activities lock-down and social distancing. In particular, we model the effect of the restrictions as *β*(t) = *β*_0_*e^−ωt^*. The choice of a decreasing exponential function is justified by the gradual application of rules, and by the increasing awareness of citizens. All these precautions will reduce effectively the basic reproduction number at each interaction.

This time dependence implies that *R*(∞) = 0, as it is expected in the so-called *fade-out* phase. On the contrary, no time dependence for *γ* is required, being the recovery time independent of the restrictions. The hypothesis *γ* = *const* could not be generally true, because of the natural progress in the treatment of patients related to the outcome of different studies and strategies adopted by the health-care system.

We notice that other studies on a time-dependent SIR model have been recently made [5]. Yet, the approach elaborated is different than what proposed in this work.

### 2.2 Choice of Free Parameters and Fit Strategy

We extract data for the compartments *I* and *R* from [6]. In order to determine the evolution of the epidemic we use the *Least Squares Method* by minimizing the following function:

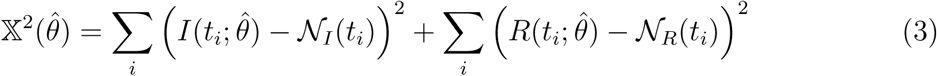

where 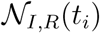 are input values for a given time, *t_i_*, and 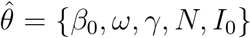 defines the set of free parameters to be determined from the data. The choice of the free parameters follows the criteria listed below:

- *β*_0_ is the infection rate at the beginning of the outbreak and is related to *R*_0_ (*t*=0);
- *ω* determines the change in time of the infection;
- *γ* is the time independent recovery rate;
- *N* and *I*_0_ are the total population involved in the outbreak and the initial number of infected population respectively.

In particular, *N* and *I*_0_ cannot be easily inferred independently: the first is actually determined by the restrictions and represents the total number that in the end will be affected by the outbreak; the second could be in principle estimated at the beginning, even if with a large systematic uncertainty, being health-care systems not ready to react immediately with a defined strategy. It is worth to point out that *N*, as in our model, could be wrongly interpreted as the total susceptible population known *a priori* in a give region or country. Since it is deduced by the data themselves, *N* should be interpreted as the susceptible population that, after restrictions, will be anyhow infected. We will referred to this concept as “virtual box”. Those considerations motivate our choice of letting both *N* and *I*_0_ as free parameters, so that they can be adapted looking at the global behaviour of the epidemic evolution.

The goodness-of-fit (GoF) is calculated exploiting a Kolmogorov-Smirnov test [9, 10]. In our set of free parameters in 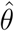*, I*(0) = *I*_0_ and *S*(0) = *N* define the initial conditions for *S* and *I*. In addition, for the chosen time dependence: *R*_0_(*t*) = *β*(*t*)/*γ*.

From the data in [6] we can also determine the number of daily new cases against time. This dependence is calculated from the result of the fit by calculating the time derivative of *I*(*t*) + *R*(*t*) + *D*(*t*), below labelled with “·” on top.

The effect of the restrictions acts basically on the *I*(*t*) trend. The curve of the infectious, or active cases, stops increasing earlier and starts decreasing with a longer tail, way more asymmetric with respect to the standard SIR prediction. The delay between *R*(*t*) and *I*(*t*), described by an unknown “transfer function”, along with the time behaviour or *R*_0_(*t*), determines the behaviour of the near and far future evolution of the epidemic.

Adding data to the fit day by day, we have been observing that the prediction is less accurate before the *I*(*t*) maximum is reached, while it is expected to become more stable and reliable after the turning point. This effect, basically determined by the fluctuation of the latest points, predict the peak time with a large uncertainty. It is worth to mention that the data points have uncertainty larger than Poissonian fluctuations, reasonably related to the methods of administration of tests and large uncertainty in locating infection sites and in taking a census of the nearest related cases.

## 3 SIRD-T: A Time Dependent SIRD Model

### 3.1 From SIR-T to SIRD-T

The SIRD-T model is a natural extension of the SIR-T when the compartments recovered (R) and deceased (D) are separated. Based on this assumption the mathematical model (1) is extended in terms of the following set of differential equations:

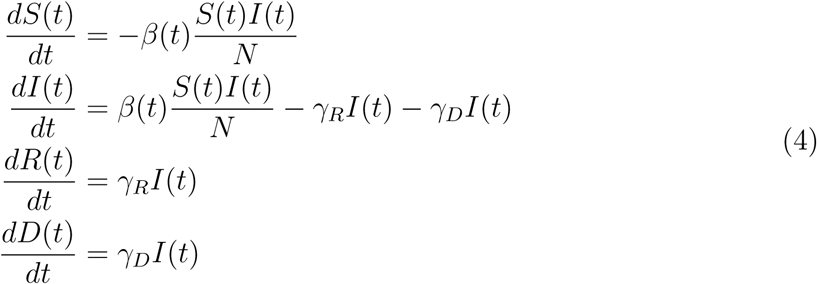

where *N* = *S*(*t*) + *I*(*t*) + *R*(*t*) + *D*(*t*) and the initial conditions are: *N*(0) = *N*_0_, *I*(0) = *I*_0_, *R*(0) = 0, and *D*(0) = 0. *β*(*t*) determines the transmission rate, *γ_R_* the recovery rate, and γ*_D_* the mortality rate. Also in this case, *R*_0_, determines the dynamic of the infection. For our specific model we define *R*_0_ = *β*/(2〈γ〉)), where (*γ*) is the average of *γ_R_* and *γ_D_*. With this definition we can write:

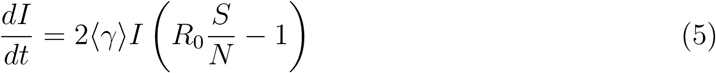

As in the SIR-T model 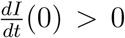 if *R*_0_ > *N*/*S*(0). Therefore, the epidemic decreases when *R*_0_ < *N*/*S*(0). Also in this model, we introduce a time dependence of *β* to account for containment measures, such as population and commercial activities lock-down and social distancing. In particular, we define *β*(*t*) = *β*_0_*e*^−^*^ωt^*, with the same meaning as before.

### 3.2 Choice of Free Parameters and Fit Strategy

We extract data for the compartments S, I, R, and D from [6]. In order to determine the evolution of the epidemic we define the following function:

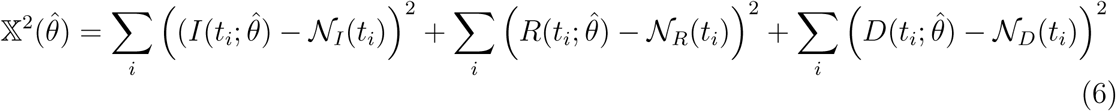

where 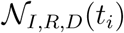 are input values for a given time, *t_i_*, and 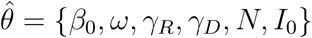 defines the set of free parameters to be determined from the data, with same meaning as in SIR-T. The differential equations in eq. (4) are solved numerically and simultaneously eq. (6) is minimised using Minuit, as described above. The GoF similarly to the previous case is calculated exploiting a Kolmogorov-Smirnov test [9, 10]. In our set of free parameters in 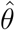, *I*(0) = *I*_0_ and *S*(0) = *N* define the initial conditions for *S* and *I*, respectively.

From the data in [6] we can also determine the number of daily new cases against time. Even in this case, this dependence is calculated from the fit result deriving the sum *I*(*t*) + *R*(*t*) + *D*(*t*).

## 4 Analysis of the Italian Data

In order to solve the system in eq. (1), while minimizing eq. (3), we exploit numerical methods. In Fig. 1 (left) we show the result of the global fit for Italian data. It turns out that 1/*β*_0_ ~ 2.3 days and 1/*γ* ~ 38 days, and *R*_0_(0) =16. The GoF is calculated to be of the order of 0.98 or greater with the KS test. Therefore, the fit reproduces well the time dependence including lock-down effects. *R*_0_ as a function of time is shown in Fig. 1 (right). Errors on the fit parameters are very small and are not reported here because we believe they are not significant due to unknown uncertainties introduced by lock-down procedures and administration of medical tests. Yet, in Fig. 2 we show an error band taking into account two standard deviations determined from the fit residuals. From this figure we can deduce: 1/*β*_0_ = 2.35 ± 0.02 days and 1/*γ* = 38.2 ± 0.7 days.

**Figure 1:**
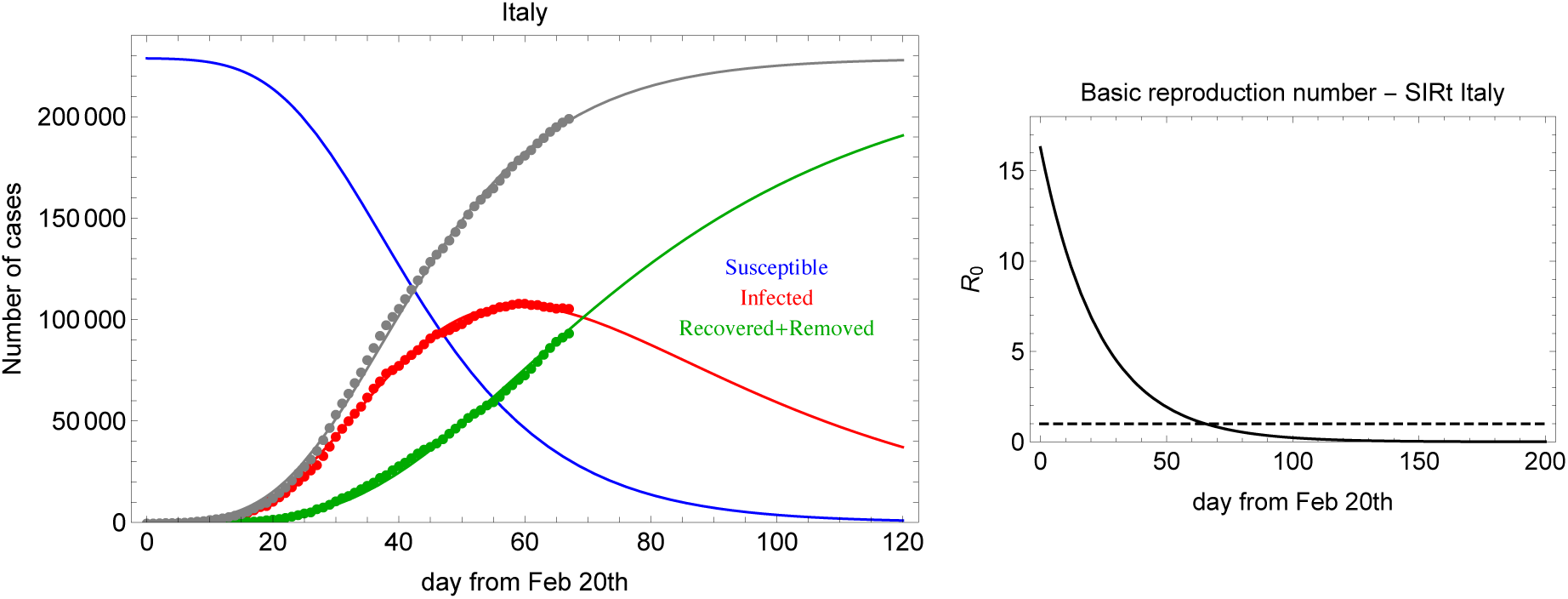
*Left:* Global fit of *I* (infected) and *R* (recovered+deceased) categories for Italian data since February 20th, 2020. *Right: R*_0_(*t*) in the SIR-T model for Italian data.

**Figure 2:**
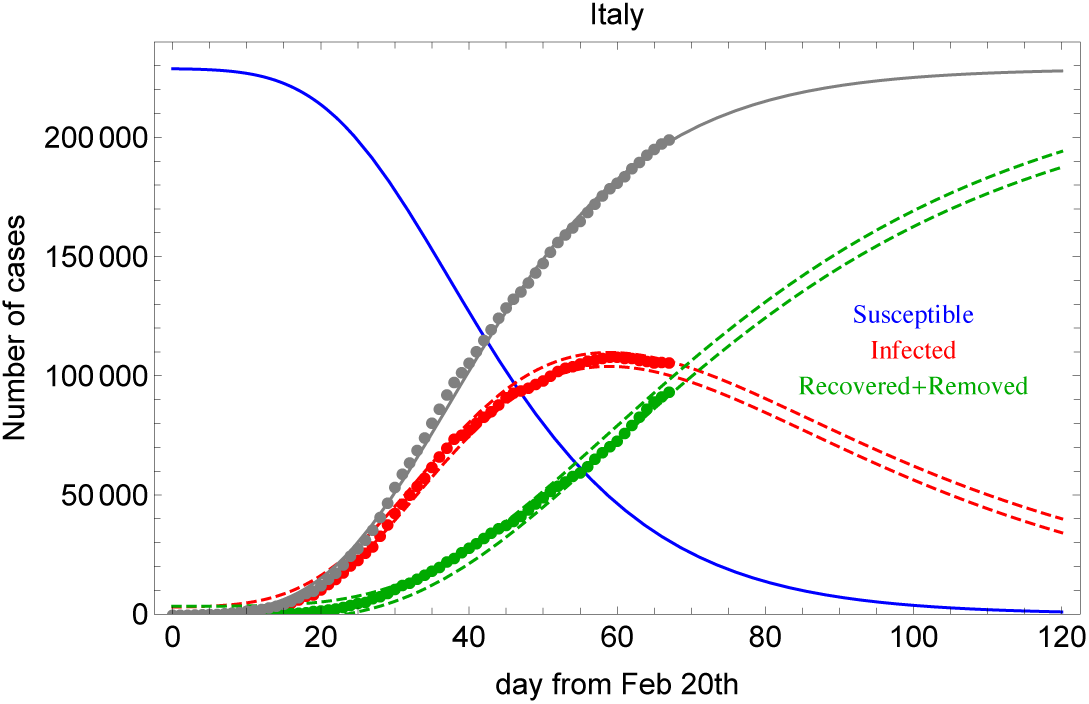
Global fit of I (infected) and R (recovered+deceased) categories for Italian data reporting error bands. See text for details.

In Fig. 3 we show the daily new and recovered+deceased cases calculated from 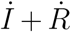 and 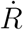, respectively. The SIR-T adopted model reproduces the time dependence of the data. This is not the case at the same statistical significance level for the SIRD-T model in eq. (4). In Fig. 4 we show the fit result for SIRD-T. As it is shown the *D* category is not described properly although the other categories are described better. For this fit it turns out that 1/*β*_0_ ~ 2.2 days and *R*_0_(0)=9. The poorer description of the SIRD-T model is basically due to the fact the sum *D*(*t*) + *R*(*t*) (simply called *R*(*t*) in the SIR-T model) is way more stable than the single compartments *D*(*t*) and *R*(*t*). This is clearly visible by comparing data from different countries, especially at the beginning of the pandemic spread, before the health-care system reaches a stable regime in classification and treatments of different symptomatologies.

**Figure 3:**
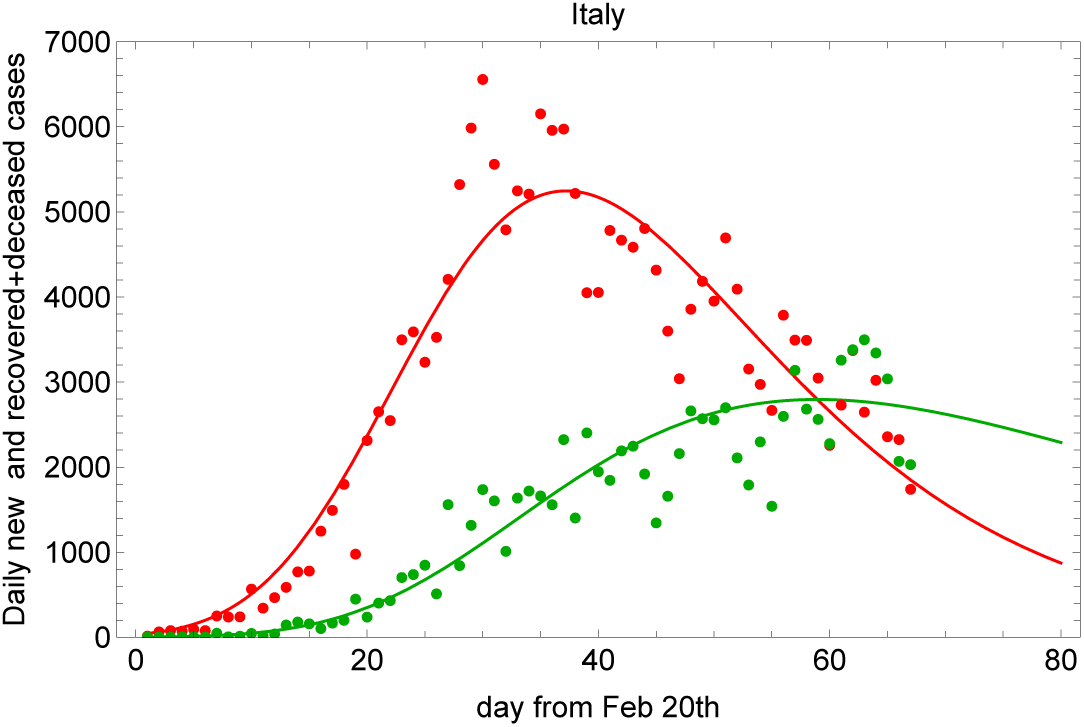
Fit to the daily new and recovered+deceased cases using 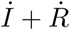 (red) and 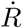 (green) for Italian data since February 20th, 2020.

**Figure 4:**
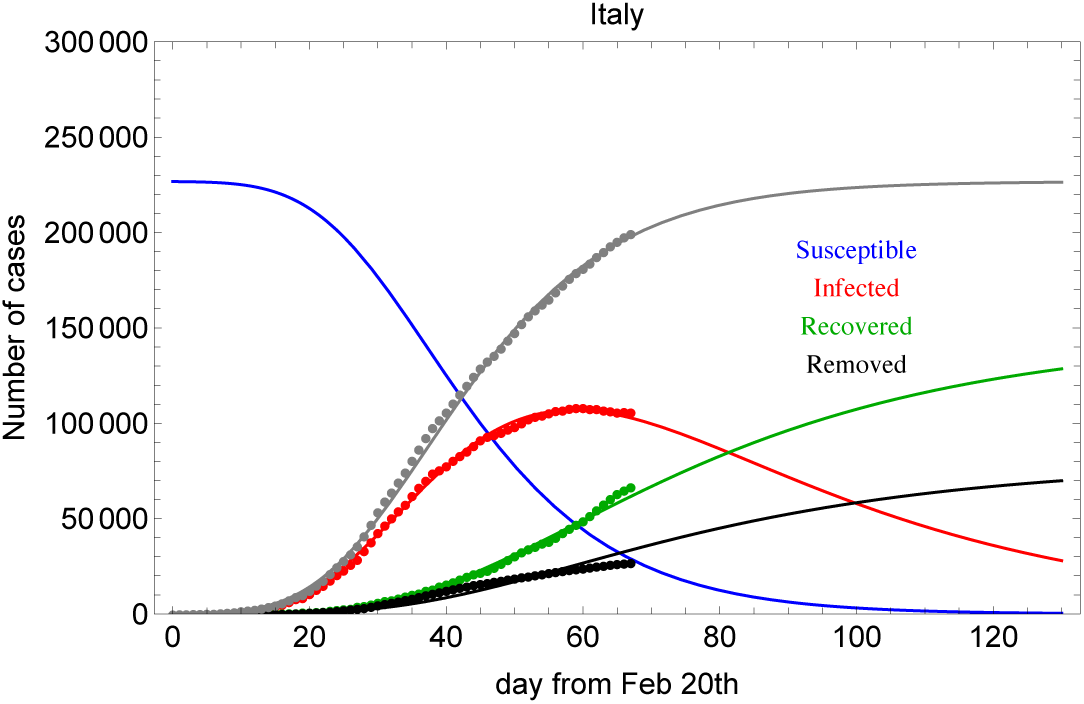
Fit with the SIRD-T model for Italian data since February 20th, 2020.

The *D*(*t*)/(*R*(*t*) + *D*(*t*)) ratio tends to the final so-called *fatality rate*, known with high accuracy only after the epidemic fade-out. As visible from [6] this ratio becomes smaller and smaller as the epidemic is close to the end in a specific country or region, as for example in China and South Korea, and/or in which the health-care system intervention is approached with a more solid strategy, as for example in Germany.

For the reasons discussed above, we can conclude that SIRD-T can improve the understanding of the epidemic spread, as compared with SIR-T, only if *R*(*t*) and *D*(*t*) show smooth diverging rates and the fatality rate decreases monotonically. In this case the different *γ_R,D_*’s give a more accurate understanding of the hidden transfer functions between *I*(*t*) and *R*(*t*), or *D*(*t*).

## 5 Analysis of Data from Other Countries

In order to compare the results above with the COVID-19 outbreak in other countries we take into account data from Spain [6]. In Fig. 5 (left) and Fig. 5 (right) we show the fit result for the SIR-T model and *R*_0_ as a function of time. For the Spanish SIR-T model it turns out that 1/*β*_0_ ~ 1.7 days, 1/γ ~ 18 days, and *R*_0_(0) =11. So, in spite of the fact that the same model works well for both sets of data, the parameters are similar (same order) but not equal within the fit errors. This could be due to the different lockdown procedure, response, and health treatment. At the same time it could indicate the limited accuracy of such a model to attempt to describe the same phenomenon in different frameworks. In Fig. 6 we show the daily new and recovered+deceased cases for Spain. In comparison with Fig. 3 the rate of change for recovered+deceased is clearly higher.

**Figure 5:**
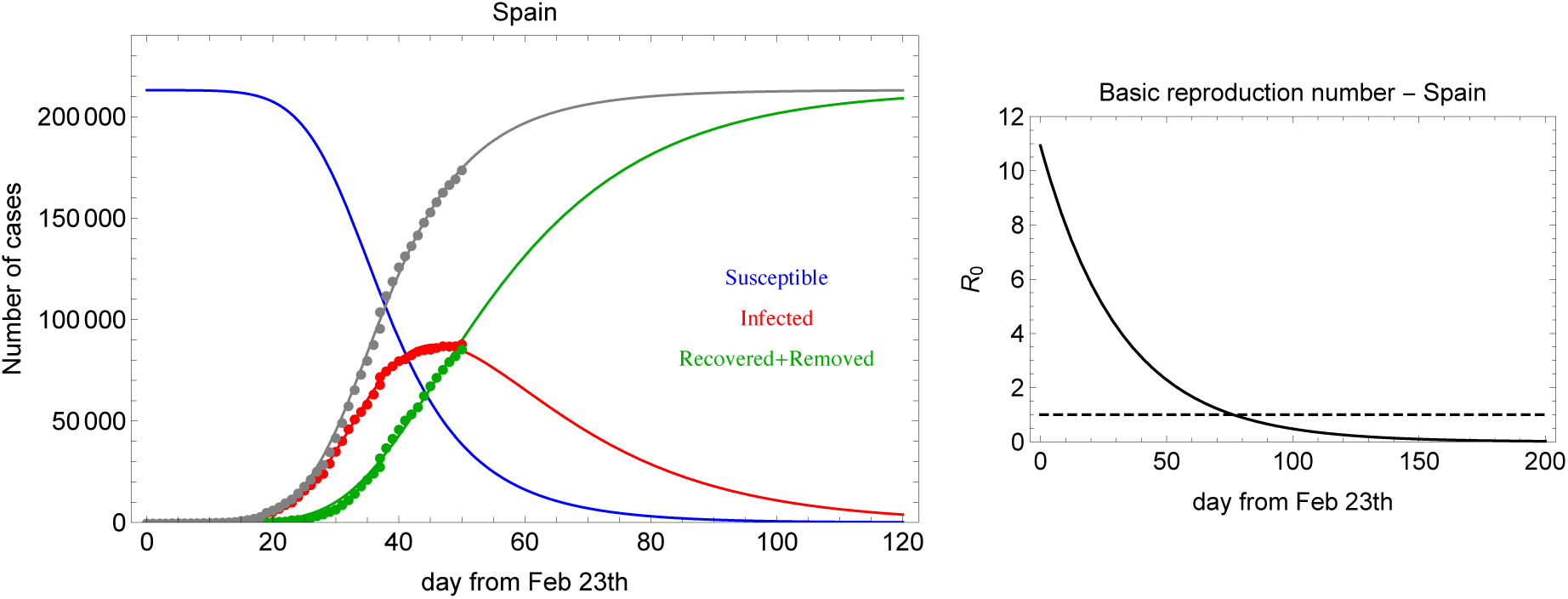
*Left:* Global fit of *I* (infected) and *R* (recovered+deceased) categories for Spanish data since February 23rd, 2020. *Right*: *R*_0_(*t*) in the SIR-T model for Spanish data.

**Figure 6:**
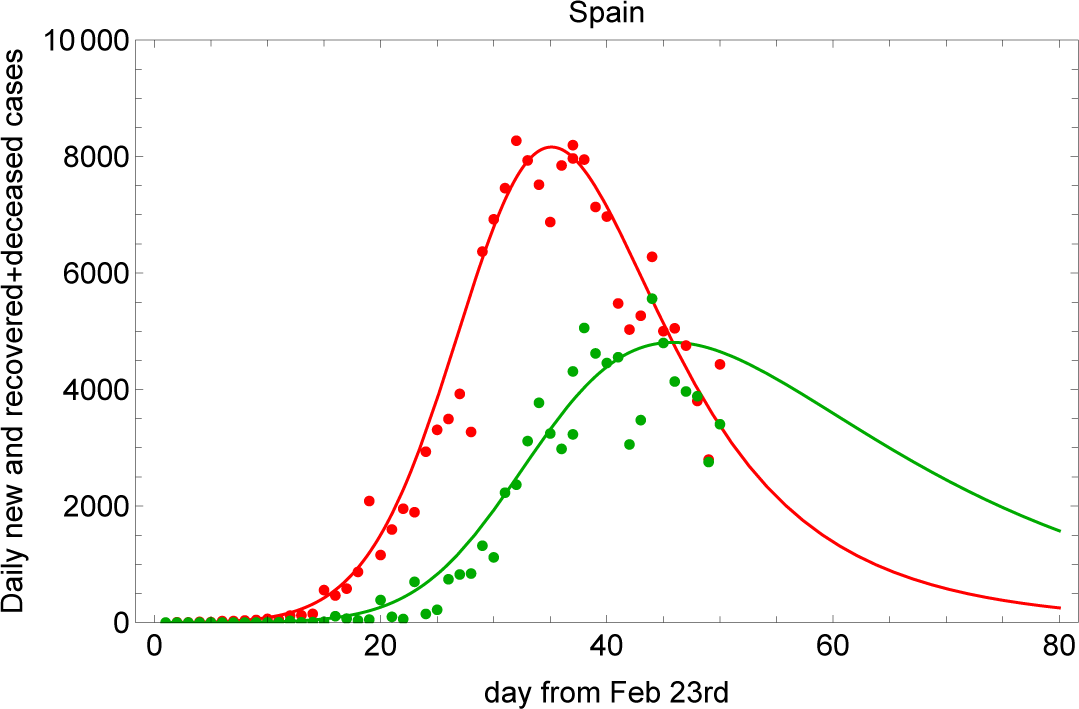
Fit to the daily new and recovered+deceased cases using 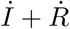(red) and 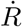 (green) for Spanish data since February 23rd, 2020.

## 6 Accounting for Asymptomatic Cases: The SIAR-T Model

In this Section we account for asymptomatic cases in the basic SIR model. The set of equations are modified as follows:

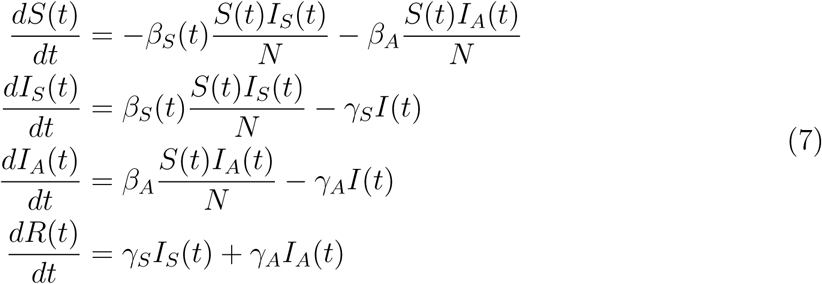

where *I_A_* accounts for the asymptomatic population and *β_S_*(*t*) = *β*_0_*e*^−^*^ωt^*. In this model to account for the lock-down effect we assume as above a transmission rate for symtomatics changing with time. On the contrary, we assume a transmission rate constant in time for undetected asymptomatics. Assuming an early exponential growth for both *I_S_* and *I_A_* we obtain the constraint: *β_S_* > *β_A_* and *γ_S_* > *γ_A_*. The fit has seven free parameters, namely 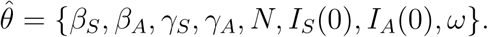 In Fig. 7 we show the result of the fit. The GoF is equal to 0.96. In addition, 1/*β*_0_ ~ 4.6 days, 1/*β_A_* ~ 6.8 days, and *R*_0_ = *β*_0_/*γ_S_* ~ 9.0. We notice that the asymptomatic population seems negligible, however, with a longer transmission time.

**Figure 7:**
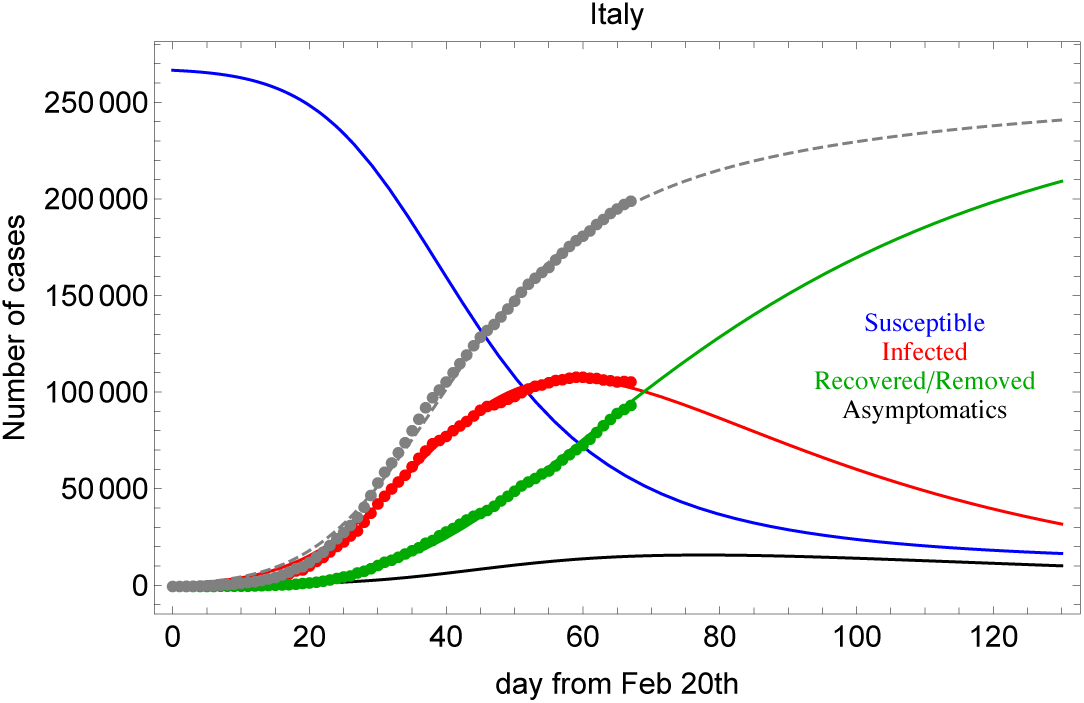
Fit to Italian data with the SIARt model.

The real motivation of this result is that the model described in (7) is sensitive only to non-constant ratio *I_S_*(*t*)/*I_A_*(*t*). If asymptomatic cases are a constant fraction of the visible cases, the model is not able to detect any anomaly in the observable compartments. As a consequence, the *I_A_*(*t*) must be interpreted as a possible deviation for the constant ratio.

It is interesting to notice that, the unknown fraction of asymptomatic cases does not invalid the SIR model and its modifications proposed in this work. This hidden component changes of course the interpretation of *R*_0_ and of other parameters that are referred to the observables.

## 7 Conclusions

In this work we proposed time dependent modifications of the SIR compartmental model, referred to as SIR-T, SIRD-T and SIAR-T, as fit models for understanding the evolution of the COVID-19 outbreaks in Italy and other countries (Spain in our case). In particular, those models allow us to infer the most important epidemic parameters fitting simultaneously active and concluded cases in presence of lock-down and social distancing restrictions.

The study that we have reported in this paper shows that the SIR model, with the changes proposed above, describes so far satisfactorily the COVID-19 Italian data time dependence with lock-down effects. The same model and assumptions work for other country data, as e.g. for Spain. Yet, in some cases reported, as an example, for the latest Spanish data, South Korean data, and Chinese data [6] some non-statistical effect are evident. These effects can be due to a strong lock-down procedure impact or to a sudden increase of the infection. In these cases the statistical model discussed in this work is not appropriate to account for important discontinuities in the data. However, it is worth mentioning that indeed the effect of the lock-down is evident in the data that we have analysed. As a matter of fact, data show small discontinuities due to the lock-down which are taken care by the model described in this work.

Among all candidates, the SIR-T model represents, given its simplicity, the best model fit, being the concluded cases (recovered+deceased) more stable with respect to the separate components, even in presence of an asymptomatic hidden component (especially if in constant ratio with the visible components). This model represents a good candidate for predicting the time evolution of the outbreak, with a sizable time scale, especially after the active case peak is overtaken.

The model has satisfactory agreement with data if restrictions play important role in limiting the possible susceptible population inside the virtual box. If restrictions are reasonably respected by people, the model predicts the end of the critical period in less than a few months. Inattention and carelessness by communities and governments will open the virtual box again exposing more people to the infection and creating a new starting point for the new cases evolution. As a consequence, an out-of-control spread of the epidemic can make the final number of cases a sizeable fraction of the total country population, quantified in many tens of millions. In cases of lack of attention to the lock-down the model fails to predict the evolution because of uncontrolled effects.

## Data Availability

https://www.worldometers.info/coronavirus/

## Acknowledgments

We would like to thank Mauro Mezzetto for introducing our discussion in the INFN COVIDSTAT project [11]. We would like to thank also Sandra Parlati for pointing out the importance of *D*(*t*) and *R*(*t*) separation in the model and Alberto di Cienzo for the useful discussions about the implementation of the SIR as fit model. Finally, we express our gratitude to Francesca Baldinelli who reviewed the terminology and the concepts used in the text.

## References

[1] https://www.who.int/health-topics/coronavirus/

[2] Neeltje van Doremalen et al. Aerosol and surface stability of HCoV-19 (SARS-CoV-2) compared to SARS-CoV-1. The New England Journal of Medicine, doi: 10.1056/NEJMc2004973.

[3] E. Lazzo et al. Suppression of COVID-19 outbreak in the municipality of Vo’, Italy.

[4] W.O. Kermack and A.G. McKendrick, Royal Society of London Proceedings Series A, 115 (1927), 700–721.

[5] Yi-Cheng Chen et al., arXiv:2003.00122v4 [q-bio.PE]9 Apr 2020.

[6] https://www.worldometers.info/coronavirus/.

[7] C. Runge, Ueber die numerische Auflösung von Differentialgleichungen. Math. Ann., 46 (1895) pp. 167–178; W. Kutta, *Beitrag zur naherungsweisen Integration von Differentialgleichungen* Z. Math. und Phys., 46 (1901) pp. 435–453.

[8] https://root.cern.ch/download/minuit.pdf.

[9] N. Smirnov, The Annals of Mathematical Statistics, Vol. 19, No. 2 (Jun., 1948), pp. 279–281.

[10] B.P. Roe Probability and Statistics in Experimental Physics, Springer-Verlag New York, Inc. 2001.

[11] https://covid19.infn.it/

